# Change in deep brain stimulation effect in Parkinson’s disease after replacement with a new generation neurostimulator

**DOI:** 10.64898/2026.05.01.26352067

**Authors:** Eline A.M.Y. Rouleau, Saskia van der Gaag, Bart J. Keulen, Marije N. Scholten, Martijn Beudel, Jolien M. ten Kate, Sabine J.E. Verkaart, Mark L. Kuijf, Marleen C. Tjepkema-Cloostermans, Esther van Veen, Eva M. de Ronde, Rianne A.J. Esselink, Erik W. van Zwet, Carel F.E. Hoffmann, Thomas A. van Essen, Niels A. van der Gaag, Rodi Zutt, M. Fiorella Contarino

**Affiliations:** Department of Neurology, Haga Teaching Hospital, The Hague, The Netherlands; Department of Neurology, Leiden University Medical Center, Leiden, The Netherlands; Department of Neurology, Amsterdam University Medical Center, Amsterdam Neuroscience, University of Amsterdam, Amsterdam, The Netherlands; Department of Neurology, University Medical Center Groningen, Groningen, The Netherlands; Department of Neurosurgery, Maastricht University Medical Center, Maastricht, The Netherlands; Department of Neurology, Maastricht University Medical Center, Maastricht, The Netherlands; Department of Neurology, Medisch Spectrum Twente, Enschede, The Netherlands; Department of Neurosurgery, Radboud University Medical Center, Donders Institute for Brain, Cognition and Behaviour, Nijmegen, The Netherlands; Department of Neurology, Radboud University Medical Center, Donders Institute for Brain, Cognition and Behaviour, Nijmegen, The Netherlands; Department of Biomedical Data Sciences, Leiden University Medical Center, Leiden, The Netherlands; University Neurosurgical Center Holland, Leiden University Medical Center, Haaglanden Medical Center and Haga Teaching Hospital, Leiden and The Hague, The Netherlands; Department of Neurosurgery, Haga Teaching Hospital, The Hague, The Netherlands; Department of Neurosurgery, Leiden University Medical Center, Leiden, The Netherlands

**Author notes:** Corresponding author: M. Fiorella Contarino.

## Abstract

Parkinson’s disease patients may experience a different therapeutic effect after replacement of the Medtronic Activa® deep brain stimulation neurostimulator with the newer Percept™ model, which features multiple independent current sources and constant-current control. We analyzed patient-reported therapeutic effect changes after Activa®-to-Percept™ replacements (AP, n=52) across six Dutch DBS-centers, comparing appropriate (AP+, n=36) and inappropriate/no (AP−, n=16) use of the manufacturer’s *replacement workflow*. Previous Activa®-to-Activa® replacements (AA, n=69) were used as reference. Worsened therapeutic effect was reported in 75.0% of AP−, 44.4% of AP+, and 21.7% of AA replacements (p<0.001). In the AP group, most patients with worsened effect were previously programmed with constant-voltage. Concluding, the risk of worsened therapeutic effect following AP replacements is higher compared to AA replacements, in particular when the *replacement workflow* is not properly used or in complex electrode configurations. We advise to use the workflow, inform the patient and plan closer follow-up appointments.

## Introduction

Deep brain stimulation of the subthalamic nucleus (STN-DBS) is a widely used treatment in the advanced stage of Parkinson’s disease (PD). Different studies show a decrease in severity of motor symptoms and shorter duration of the off-drugs phase and a decrease of dyskinesia ^1^. Furthermore, it improves quality of life ^2^. DBS can induce reversible side effects like dysarthria, paresthesia, eye movement abnormalities, deterioration in balance or psychiatric symptoms due to overstimulation of adjacent structures ^3^. These side effects can be treated by adjusting the stimulation parameters.

In 2020, a new model of implantable pulse generator (IPG), Percept™ (Medtronic Inc.) became available and DBS centers gradually started to replace the previously implanted Activa® (Medtronic Inc.) IPGs with the new model, a process that is still ongoing worldwide.

While the Activa® is a single source system which can be programmed in constant voltage (CV) or constant current (CC) mode, the Percept™ works with multiple independent current sources ^4^, and can only be programmed in CC mode ^5^. On the Activa®, more advanced stimulation configurations (other than single monopolar or bipolar), could only be programmed in CV. With CV mode, the delivered current is dependent on the impedance and could be less stable and unevenly distributed across the multiple active contacts. With CC mode and multiple current sources, the delivered current is independent from the electrodes’ impedance and can be defined independently for each electrode ^5^. To assist the conversion of the stimulation parameters from one system to the other, Medtronic introduced a *replacement workflow* ^6^. In the case of CV settings, this is based on the recorded therapeutic current or conversion to current using Ohms law (I (current) = V (voltage) / R (resistance)). With CC settings, the amplitude or applied current can be directly read from the stimulation screen ^6^. However, it is a common impression among the clinicians that the new parameters do not always result in a comparable therapeutic effect. This yields discomfort for the patients, sometimes with potentially dangerous situations (e.g., falling) as a result, and increases the need for medical care.

Previous studies regarding the switch from CV to CC stimulation did not show any differences in motor- or non-motor symptoms although these studies do describe the need of adjustment of initial DBS settings in some cases ^7–9^. These studies evaluated changing the mode of stimulation in the same IPG (Activa®, Medtronic Inc.) ^10,11^ or changing mode of stimulation because of the introduction of mixed implants (i.e., combining leads from one manufacturer to an IPG from another) ^8,9,12–14^.

The goal of this multicenter retrospective study is to review the clinical results after replacing Activa® with Percept™ in a cohort of patients with PD and STN-DBS in order to analyze the extent of the issue and the possible predisposing factors, in order to improve this process.

## Results

### Patient population

A total of 88 patients with PD and STN-DBS who had an Activa® to Percept™ (AP) replacement were considered for inclusion. Of these, 8 were not included because of unstable settings before replacement, resulting in 80 included patients. In only one center (Amsterdam University Medical Center (Amsterdam UMC)) more than 25 replacements were performed in the selected period but not included due to the predefined limit. In order to include 80 Activa® to Activa® (AA) replacements, 92 consecutive patients were screened: of these 12 could not be included (one without informed consent; 11 with unstable settings) (supplementary table 1). After initial screening, three patients in the AP group were secondarily excluded because the IPG had reached *end of service* prior to replacement (n=1) or because a different target than the STN was used (n=2). In addition, 11 more patients in the AA group and three patients in the AP group for which the therapeutic effect after IPG replacement was unknown were excluded (supplementary table 1). This resulted in 69 patients in the AA control group and 74 patients in the total AP group. Among these 74 patients, in 36 the *replacement workflow* was appropriately used (AP+), in 16 the *replacement workflow* was not or inappropriately used (AP−) and for 22 it was unknown whether the *replacement workflow* was used (AP?).

In the AA group, 10/69 (14.5%) patients had an IPG replacement to an Activa® RC. In the AP groups all IPGs were replaced with a Percept*™* PC.

The baseline patient characteristics and stimulation parameters of each of the four (sub)groups (AA, AP+, AP− and AP?) are reported in table 1. Between the groups, there was a difference in duration of DBS-treatment (p=0.009) and pulse width (p=0.043). However, pairwise comparisons with adjusted p-values showed no statistically significant differences between the individual groups. Furthermore, there was a difference in stimulation configuration (CC/CV) between groups (p=0.005).

**Table 1.**
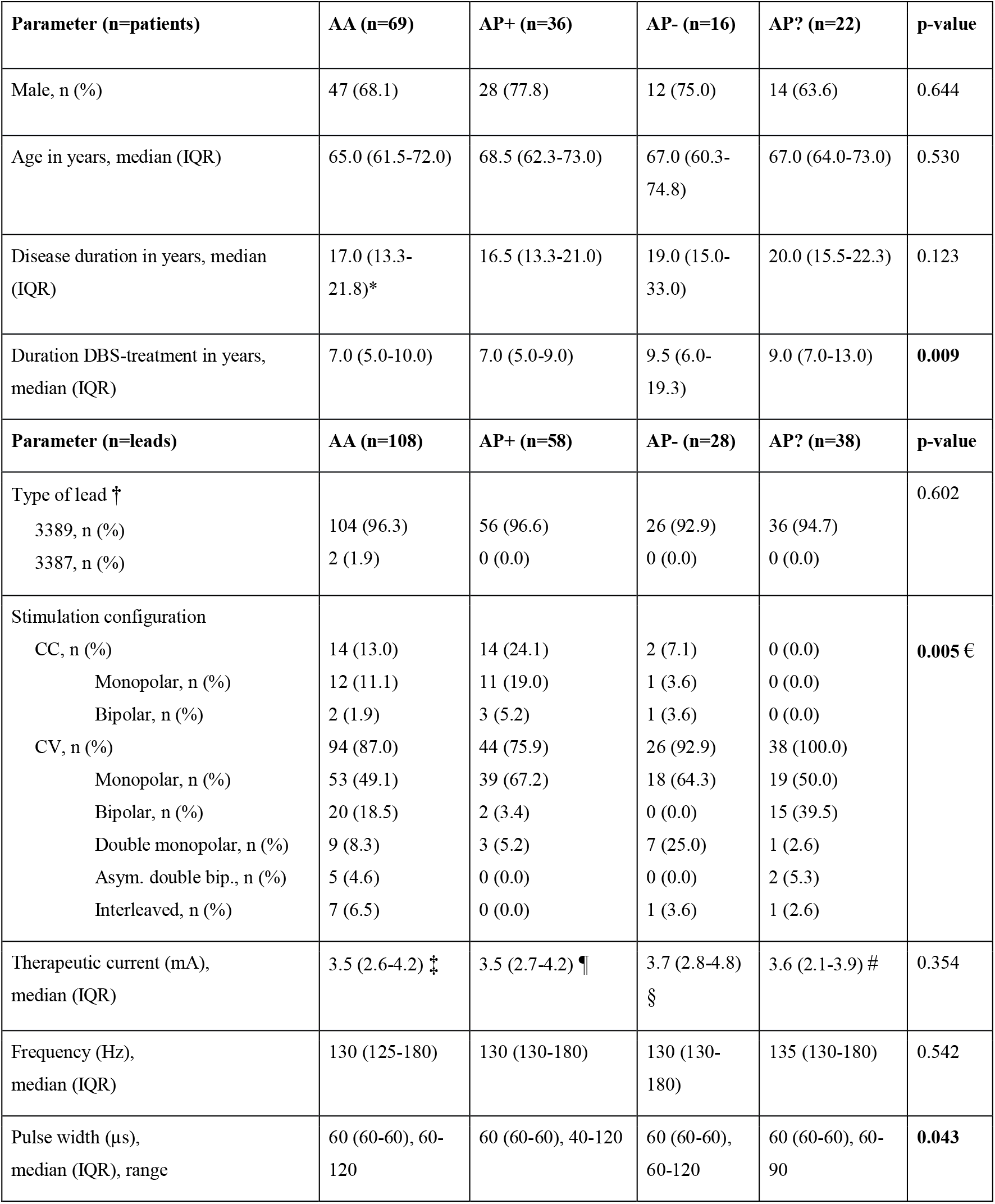
Patient characteristics per (sub)group and DBS characteristics per lead on Activa® IPG before replacement. Leads with known therapeutic effect: 108/138 AA, 58/72 AP+, 28/32 AP−, 38/44 AP?. * = Available for 68/69 patients, † Missing for two leads in every (sub)group, € Result for distribution of total CC and CV between groups, ‡ Available for 79 leads, ¶ Available for 48 leads, § Available for 24 leads. # Available for 22 leads. AA= Activa® to Activa® replacement, AP+ = Activa® to Percept™ using *replacement workflow*, AP– = Activa® to Percept™ not/inappropriately using *replacement workflow*, AP? = Activa® to Percept™ where it was unknown whether *replacement workflow* was used. IPG= implantable pulse generator, IQR = interquartile range.

| Parameter (n=patients) | AA (n=69) | AP+ (n=36) | AP- (n=16) | AP? (n=22) | p-value |
| --- | --- | --- | --- | --- | --- |
| Male, n (%) | 47 (68.1) | 28 (77.8) | 12 (75.0) | 14 (63.6) | 0.644 |
| Age in years, median (IQR) | 65.0 (61.5-72.0) | 68.5 (62.3-73.0) | 67.0 (60.3-74.8) | 67.0 (64.0-73.0) | 0.530 |
| Disease duration in years, median (IQR) | 17.0 (13.3-21.8)* | 16.5 (13.3-21.0) | 19.0 (15.0-33.0) | 20.0 (15.5-22.3) | 0.123 |
| Duration DBS-treatment in years, median (IQR) | 7.0 (5.0-10.0) | 7.0 (5.0-9.0) | 9.5 (6.0-19.3) | 9.0 (7.0-13.0) | **0.009** |
| Parameter (n=leads) | AA (n=108) | AP+ (n=58) | AP- (n=28) | AP? (n=38) | p-value |
| Type of lead †<br>3389, n (%)<br>3387, n (%) | 104 (96.3)<br>2 (1.9) | 56 (96.6)<br>0 (0.0) | 26 (92.9)<br>0 (0.0) | 36 (94.7)<br>0 (0.0) | 0.602 |
| Stimulation configuration<br>CC, n (%)<br>Monopolar, n (%)<br>Bipolar, n (%)<br>CV, n (%)<br>Monopolar, n (%)<br>Bipolar, n (%)<br>Double monopolar, n (%)<br>Asym. double bip., n (%)<br>Interleaved, n (%) | 14 (13.0)<br>12 (11.1)<br>2 (1.9)<br>94 (87.0)<br>53 (49.1)<br>20 (18.5)<br>9 (8.3)<br>5 (4.6)<br>7 (6.5) | 14 (24.1)<br>11 (19.0)<br>3 (5.2)<br>44 (75.9)<br>39 (67.2)<br>2 (3.4)<br>3 (5.2)<br>0 (0.0)<br>0 (0.0) | 2 (7.1)<br>1 (3.6)<br>1 (3.6)<br>26 (92.9)<br>18 (64.3)<br>0 (0.0)<br>7 (25.0)<br>0 (0.0)<br>1 (3.6) | 0 (0.0)<br>0 (0.0)<br>0 (0.0)<br>38 (100.0)<br>19 (50.0)<br>15 (39.5)<br>1 (2.6)<br>2 (5.3)<br>1 (2.6) | **0.005 €** |
| Therapeutic current (mA), median (IQR) | 3.5 (2.6-4.2) ‡ | 3.5 (2.7-4.2) ¶ | 3.7 (2.8-4.8) § | 3.6 (2.1-3.9) # | 0.354 |
| Frequency (Hz), median (IQR) | 130 (125-180) | 130 (130-180) | 130 (130-180) | 135 (130-180) | 0.542 |
| Pulse width (µs), median (IQR), range | 60 (60-60), 60-120 | 60 (60-60), 40-120 | 60 (60-60), 60-120 | 60 (60-60), 60-90 | **0.043** |

No statistically significant differences in baseline patient characteristics or stimulation parameters were found between the included patients and those excluded because of unknown effect (except for current intensity, p=0.049), nor between the total AA and AP group (except for DBS-treatment duration, p=0.048) (supplementary table 2).

### Therapeutic effect and adjustment of settings after IPG replacement

In the AA group, therapeutic effect worsened in 21.7% of patients (15/69) compared to 54.1% (40/74) in the total AP group. Within the AP group, a worsened therapeutic effect was seen in 44.4% of patients (16/36) in the AP+ group and 75.0% (12/16) in the AP– group (figure 1). The overall binary logistic regression model was significant (χ^2^ (2) = 17.761, p < 0.001) and explained 18.8% of variation in therapeutic effect (Nagelkerke R^2^). The odds for having a worsened therapeutic effect after AA replacement was 0.28. The odds ratio after AP+ replacement was 2.88 (95% CI= 1.21 to 6.88) relative to the reference category (AA group), whereas after AP−replacement it was 10.75 (95% CI= 3.04 to 38.38).

**Figure 1:**
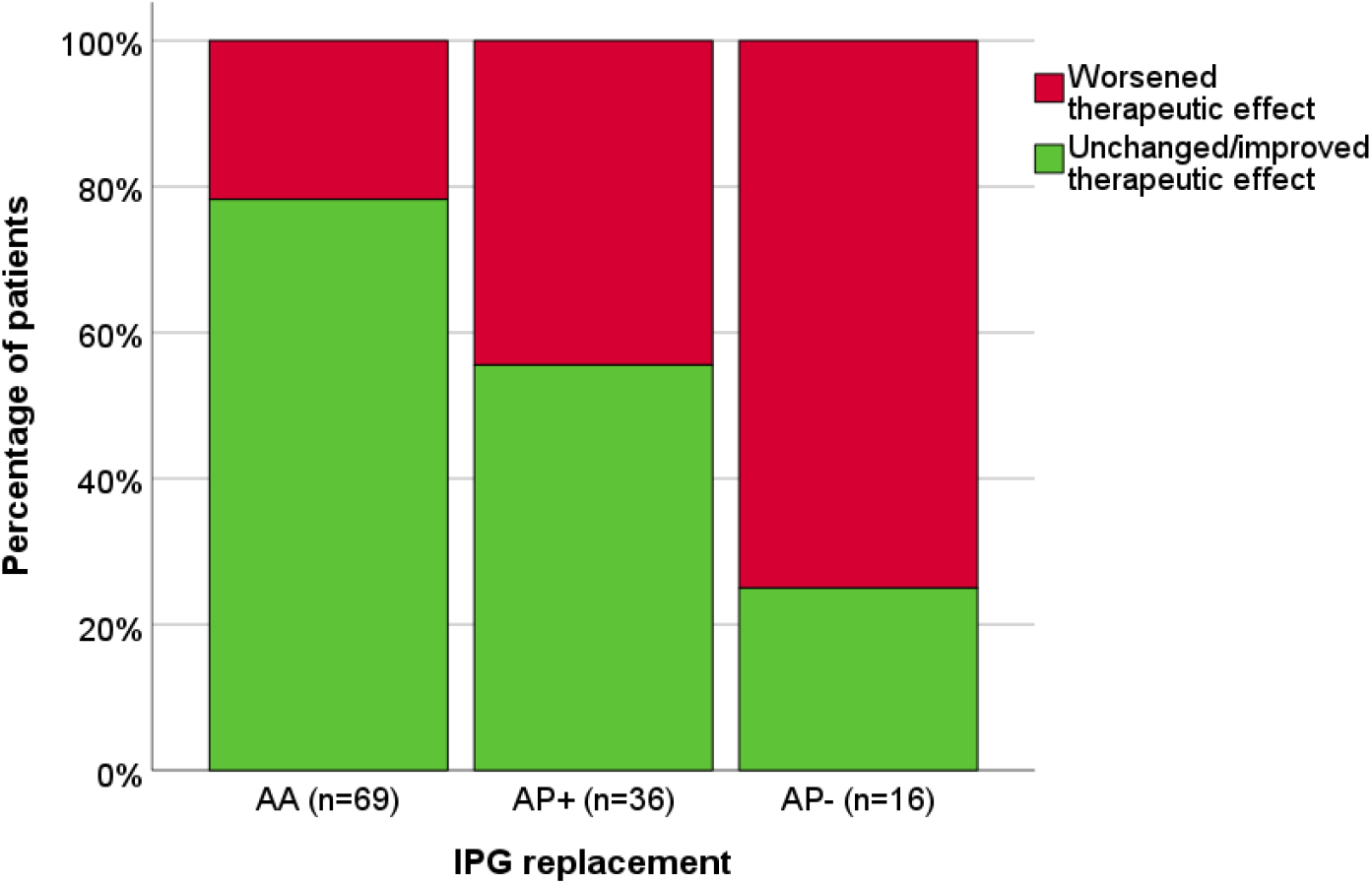
Therapeutic effect after IPG replacement per (sub)group. Replacements divided in AA (Activa® to Activa® replacements), AP+ (Activa® to Percept™ replacements using workflow) and AP– (Activa® to Percept™ replacements not/inappropriately using workflow) with corresponding therapeutic effect divided in worsened (red) and unchanged/improved (green) therapeutic effect. IPG= implantable pulse generator.

In the AP– group the workflow was either not used (11/16) or inappropriately used (5/16, double monopolar settings in all cases) (supplementary table 3).

Among the 15 patients in the AA group who worsened, 11 had adjustments of DBS settings, two spontaneously improved over time, one had medication adjustments, and one was referred to a speech therapist because of dysarthria. In the 16 patients in the AP+ group who worsened, 14 patients had their settings changed in the first three months and of the other two patients, one reported the post-replacement deterioration only later on. Lastly, in the AP– group, all 12 patients who worsened needed stimulation parameter adjustments (figure 2).

**Figure 2:**
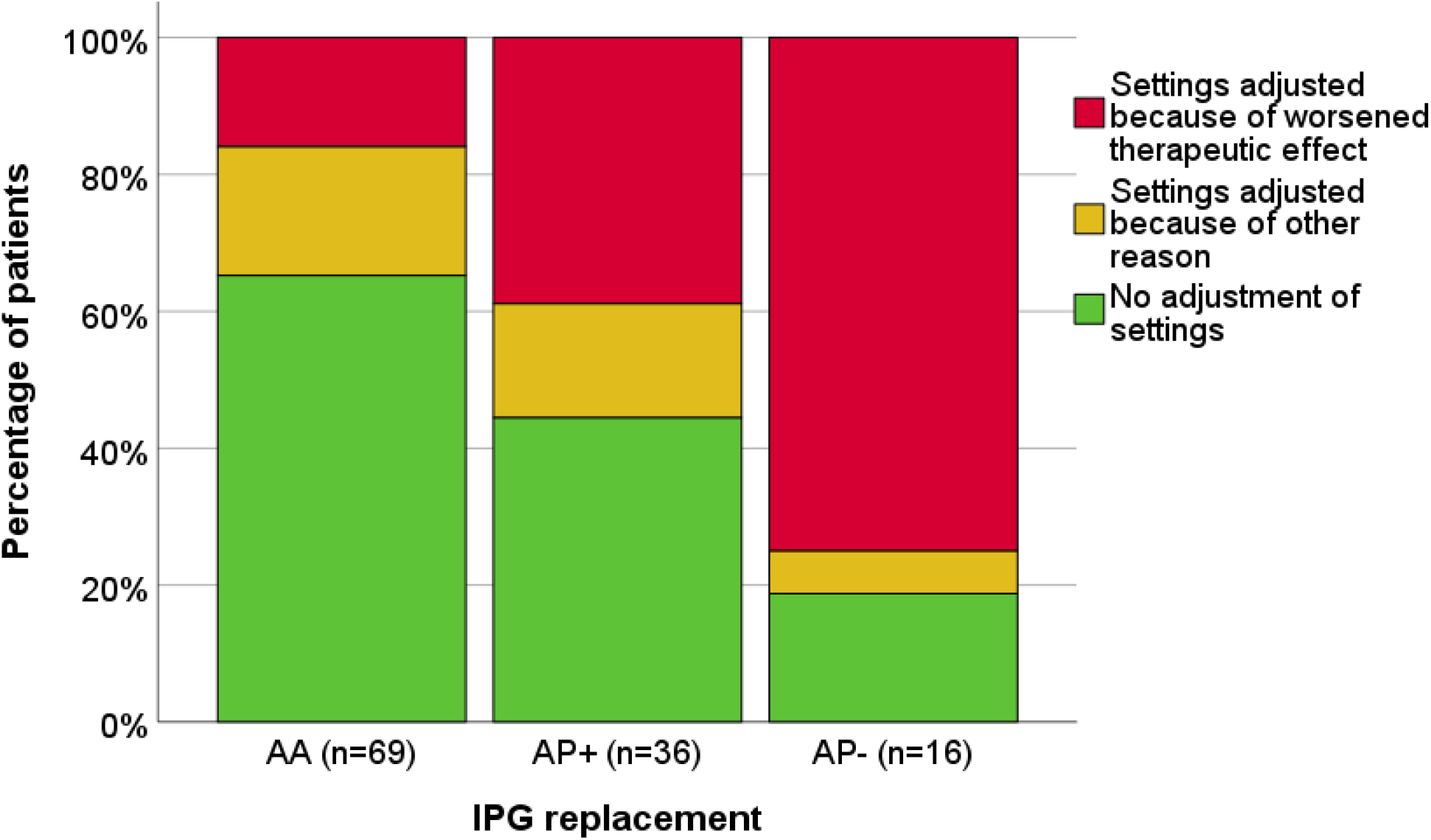
Adjustment of settings following IPG replacement. Replacements divided in AA (Activa® to Activa® replacements), AP+ (Activa® to Percept™ replacements using workflow) and AP– (Activa® to Percept™ replacements not/inappropriately using workflow) with percentage of patients needing adjustments in stimulation settings: settings adjusted because of worsened therapeutic effect (red); settings adjusted because of other reason (yellow), no adjustment of settings (green). IPG= implantable pulse generator.

The median number of adjustments made in consultation with the clinician in case of worsened effect was two (range 1-5) for the AA group, one (range 0-3; three patients independently adjusted settings) for the AP+ group, and 2 (range 1-4) for the AP– group. Some patients with worsened effect still did not reach a comparable effect after three months (AA: 2; AP+: 4; AP–: 1) or it was unknown whether a comparable effect was reached (AP_–:_ 1).

Of the 74 patients in the total AP group, 45 patients (60.8%) had at least one previous AA replacement (median 1; range 1-7). For 40 of these patients, the therapeutic effect after both the AA and AP replacements was known. Of those patients, 20 (50.0%) had a worsened therapeutic effect after AP replacement and 12 (30.0%) after AA replacement (figure 3). There was a trend towards more patients with worsened effect after AP compared to their AA replacement, although this was not statistically significant (p = 0.077) (figure 3).

**Figure 3:**
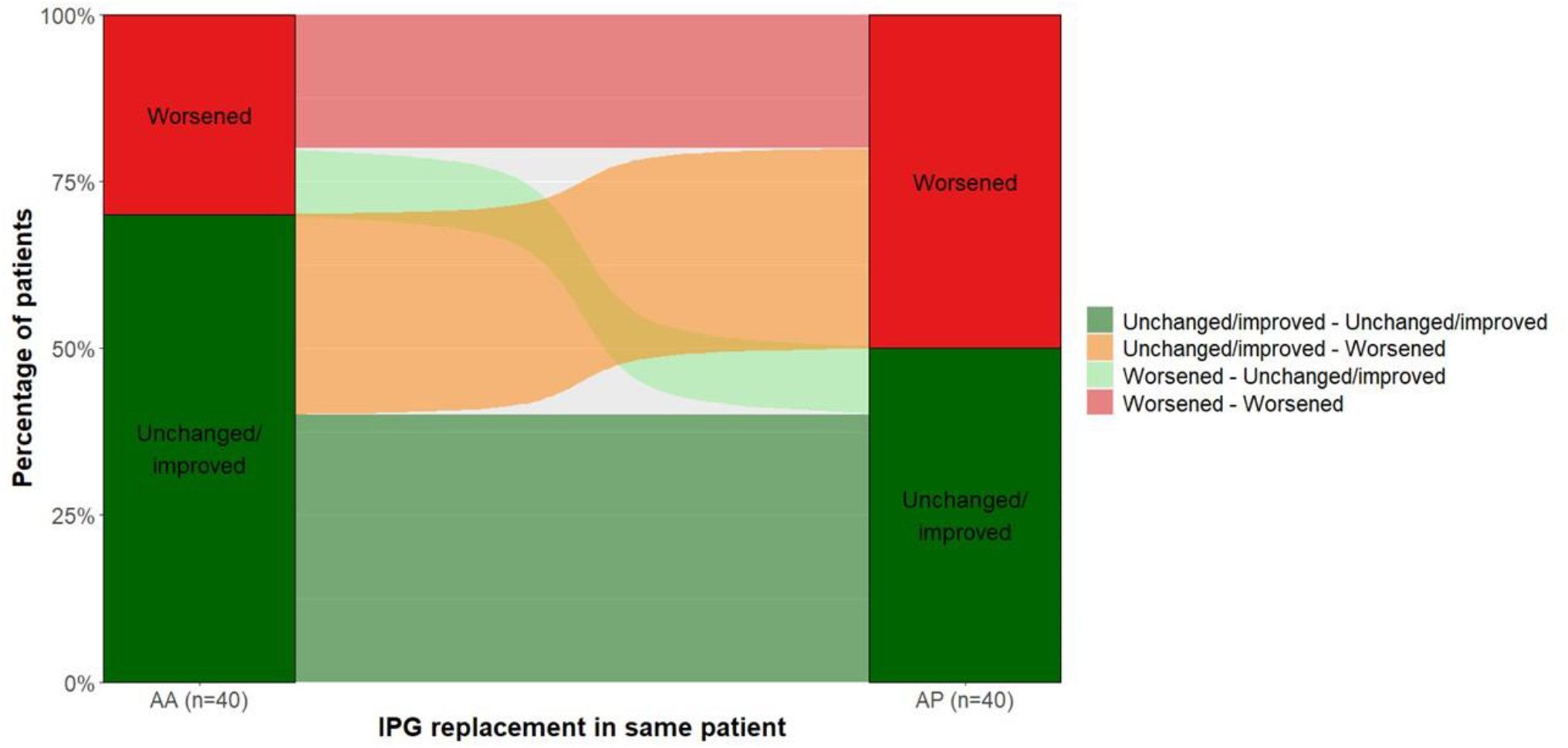
Therapeutic effect after Activa® to Percept™ and previous Activa® to Activa® replacement in the same patient. Sankey diagram of the connection between therapeutic effect for both replacements in the same patient, displayed by the flow and corresponding color. All AP replacements, regardless of use of the *replacement workflow*, were included. AA = Activa® to Activa® replacement, AP = Activa® to Percept™ replacement, IPG= implantable pulse generator.

### Specification of worsened therapeutic effect

The type of complaints (over- or understimulation) did not differ between the AA, AP+ and AP– groups (patient: χ^2^(4) = 0.735, p = 0.972, figure 4; lead: χ^2^(2) = 0.819, p = 0.712, supplementary figure 1). Of the two patients in the AA group with worsening because of a combination of symptoms, one patient showed more PD symptoms without specification of side as well as stimulation-induced dysarthria, the other patient showed an increase in tremor (side unknown) as well as stimulation-induced dysarthria and axial dyskinesia. The one patient in the AP+ group with a combination of symptoms showed more axial PD symptoms as well as more dyskinesia (side unknown). The one patient in the AP– group showed more tremor (side unknown) as well as stimulation-induced dysarthria.

**Figure 4:**
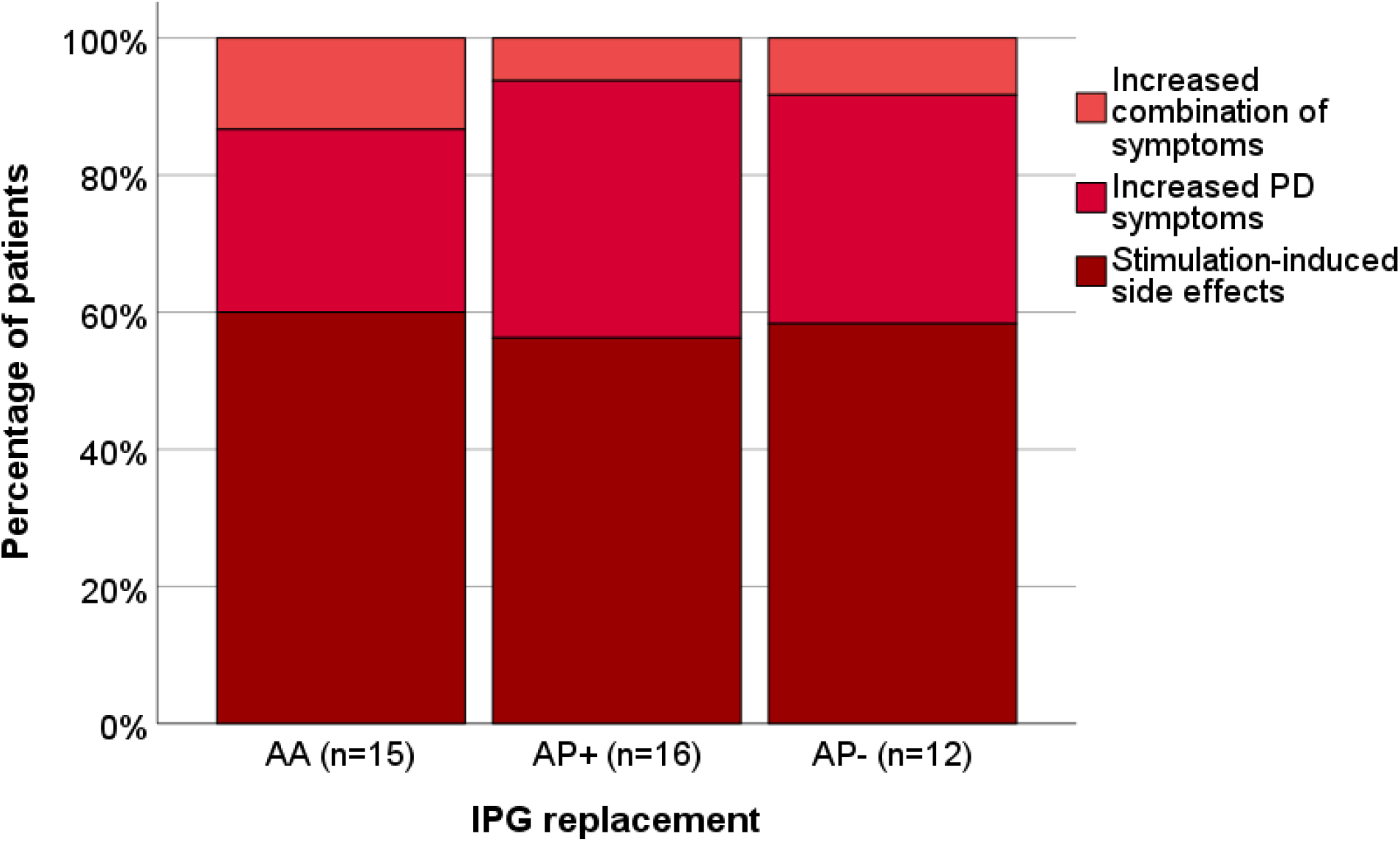
Specification of worsened therapeutic effect per patient after IPG replacement. Replacements divided in AA (Activa® to Activa® replacement), AP+ (Activa® to Percept™ replacement using workflow) and AP– (Activa® to Percept™ replacement not/inappropriately using workflow) with specification of worsened therapeutic effect: increased combination of PD symptoms and stimulation-induced side effects (orange); increased PD symptoms (understimulation, light red); stimulation-induced side effects (overstimulation, dark red). PD= Parkinson’
ss disease, IPG= implantable pulse generator.

### Stimulation configuration before IPG replacement

Before replacement, the leads were programmed in CV in the following percentages (respectively worsened vs. unchanged/improved therapeutic effect): AA (87.5% vs. 87.0%) (supplementary figure 2), AP+ (100.0% vs. 68.2%) (figure 5: panel a) and AP– (88.2% vs. 100.0%) (supplementary figure 2). Of interest, all leads programmed in CC had an unchanged/improved effect after AP+ replacement. When considering the stimulation configuration for AP versus previous AA replacement in the same patient, all leads with worsened effect were programmed in CV (figure 5: panel b), whereas 3.5% of the leads with unchanged effect were programmed in CC before AA replacement as well as 13.0% before AP replacement.

**Figure 5:**
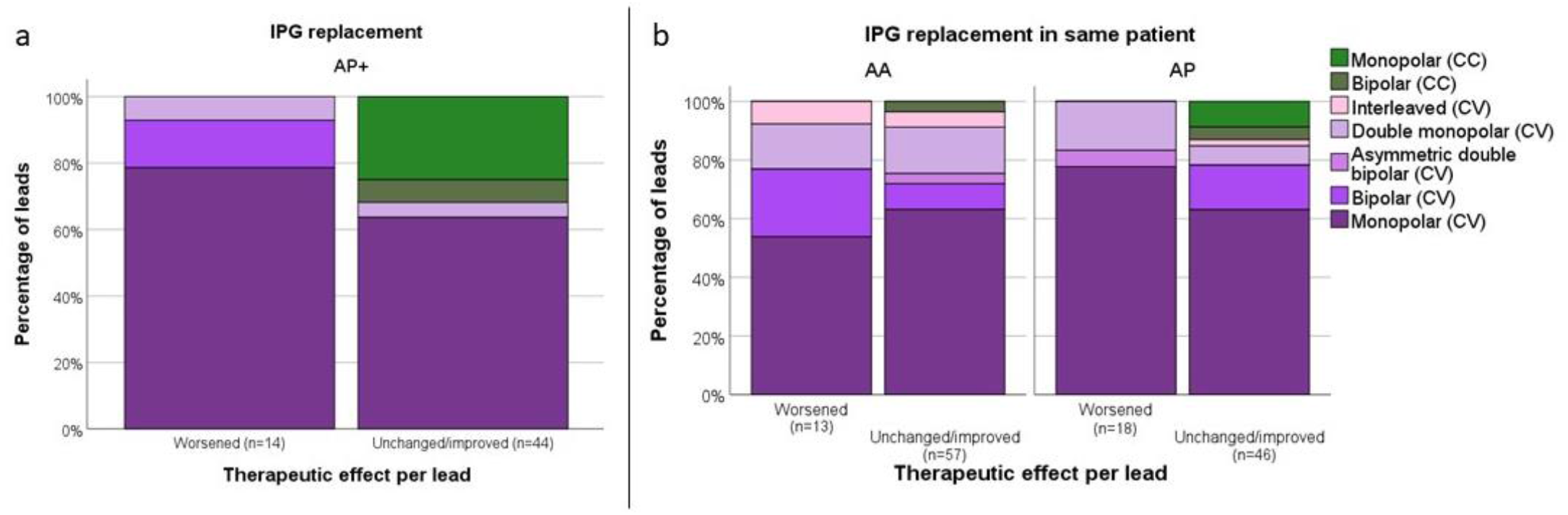
Specification of pre-replacement stimulation configuration per lead. **a:** AP+ replacements (Activa® to Percept™ replacement using workflow) divided in worsened and unchanged/improved therapeutic effect. Leads with known therapeutic effect: 58/72. **a:** replacements divided in previous AA (Activa® to Activa® replacement) and recent AP (Activa® to Percept™ replacement) in the same patient as well as in worsened or unchanged/improved therapeutic effect. All AP replacements, regardless of use of the *replacement workflow*, were included. Leads with known therapeutic effect from patients with known general effect for both replacements: AA 70/80, AP 64/80. CC = constant current, CV = constant voltage, IPG= implantable pulse generator.

## Discussion

Patients undergoing Activa® to Percept™ replacements (including AP+, AP– and AP?) were twice more likely (21.7% vs. 54.1%) to experience a worsened therapeutic effect compared to patients undergoing Activa® to Activa® replacements. This occurred when the *replacement workflow* was not or inappropriately used (AP_–:_ 75.0%), but also when it was applied appropriately (AP+: 44.4%). This suggests that the algorithm underlying the workflow may not fully resolve the challenges of transitioning to a different type of stimulator in all cases. It is plausible that the replacement workflow was increasingly used (in the right way) as experience in the different centers grew, which could mean that the number of patients belonging to the AP– group reduced over time.

Our data suggest that when translating previous CV configurations and in particular asymmetric double bipolar and double monopolar configurations, the conversion formula may be less accurate. Indeed, all leads in the AP+ group with worsened effect were programmed in CV. The algorithm assumes that the current reported in the tablet and the current calculated using Ohm’s law based on the reported impedance is equal, which might not be the case in real life settings. In addition, because of the transition from single source to multiple sources, the algorithm dictates that when multiple cathodes are active with an equal amount of cathodes and anodes on the lead, the current has to be divided among the cathodes, while with an unequal amount, it should not ^6^. This step might introduce some mismatch in effect, considering that in the previous single-source system variation in tissue impedance may have resulted in an unequal distribution of current across multiple active contacts, which is not accounted for in the algorithm. At the end of the workflow, the manufacturer advises to titrate the stimulation amplitude as needed. However, right after replacement, this step is often overlooked because the difference in therapeutic effect is not always immediately evident.

We did not observe a clear trend toward overstimulation (stimulation-induced side effects) or understimulation (more PD symptoms) in case of worsened effect (figure 4, supplementary figure 1), likely due to individual variability, so based on these data we cannot propose an adjustment in the replacement algorithm.

In our study, all replacements concerned neurostimulators from the same manufacturer. Previous studies have looked at replacing a single-source (CV) IPG with a multiple-sources (CC) IPG from another manufacturer in PD patients (in most cases Medtronic to (rechargeable) Boston Scientific® IPGs ^8,9,12,13^, or in one study to St. Jude Neuromodulation IPGs ^14^). There was a considerable heterogeneity between these studies in study design (retrospective vs. prospective), number of patients, included disorders, outcome measures and follow-up durations.

Two studies analyzed PD patients separately ^12,14^, whereas three other studies integrated patients with dystonia ^8^, essential tremor ^9^ or both ^13^ in their analysis. The pre-replacement therapeutic current recorded by the system or calculated by Ohm’s law was most frequently used to determine post-replacement DBS settings ^8,13,14^. However, in one study, impedance of the used contacts on the new device was used to calculate the ‘theoretical amplitude’ ^12^ and in another study the amplitude was intentionally reduced by 0.5 mA after replacement ^9^. No specific algorithm was used for different configurations as described for the Activa® to Percept™ replacements. While in the literature ^8,9,12–14^ prospective or retrospective analysis included stimulation adjustments and evaluation performed from three to 31 months after replacement, in our study we included only stimulation adjustments done within three months post-replacement, which makes an effect of disease progression less likely.

In three studies, UPDRS-III scores pre and post-replacement at varying follow-up durations were reported ^8,13,14^, and overall no significant changes were found. In four studies, subjective outcome measures were assessed at varying follow-up durations. A three-point Clinical Global Impression (CGI) scale on clinical efficacy of CC stimulation ^14^ and a seven-point CGI-improvement scale ^8,12^ for patients and clinicians generally suggested an unchanged therapeutic effect in the majority of patients. Another study assessed patient/caregiver satisfaction by surveys, which showed that 92% of respondents (patient or caregiver) reported no change in symptomatology since replacement, while 33% actually needed additional programming visits ^9^.

Of interest, the outcomes after replacement in the aforementioned studies were mostly assessed after adjustments had been made to the initial post-replacement settings. This was the case in one-third of patients in one study ^9^ the majority of patients in two studies ^8,13^ and in all patients in another study ^12^. Interestingly, the latter study reported that DBS settings were adjusted in only 19.3% of electrodes in the control group of patients switching to the same CV IPG ^12^. On the other hand, they reported no differences in duration of hospitalization (3 days for CV-CC vs. 2 days for CV-CV), need for rehospitalisation (5 vs. 5 patients) and total number of outpatient visits to adjust stimulation parameters during the following three months (15 vs. 22) ^12^. Another study reported a mean number of 3.3 ± 1.6 reprogramming sessions necessary to obtain the same clinical outcome, which was similar to previous CV-CV IPG replacements in the same patients ^14^.

While most studies concluded that there were no changes in therapeutic effect after IPG replacement, DBS settings had to be adjusted in a substantial amount of cases, which might imply that the initial post-replacement therapeutic effect was in fact different. This is in line with our study where in the AP– group all patients with worsened effect needed stimulation adjustments, suggesting real deterioration of effect, while in the AA group not all patients with worsened effect needed stimulation adjustments, with some experiencing spontaneous improvement, which might suggest a nocebo effect due to replacement or a temporary change in effect. The need of stimulation adjustments after replacement implies extra burden for the patients and extra use of clinical resources.

Two studies looked at switching from CV to CC on the same Activa® IPG in PD patients: in this case there was no issue of changing from a single source to a multiple sources device. In neither study differences in motor or non-motor outcomes were found. In one study no adjustments in stimulation settings were needed ^11^ while in the other study setting adjustments were performed in some cases ^10^.

Our results give ground to a very common observation among the DBS clinicians, who often perceive a mismatch in effect after replacing Activa® with Percept™ neurostimulators, and serve as a warning to the centers in the world that are now in or approaching this transition.

Among the strengths of this study are the relatively large multicenter population, the use of a control group with AA replacements to account for subjective changes unrelated to the difference in neurostimulator, and the systematic approach.

Due to its retrospective nature, this study has some limitations. We did not use an objective primary outcome measure, as we analyzed the patient’s experience of the therapeutic effect and their use of care. However, the use of subjective measures is unlikely to affect the observed differences between groups. In addition, the patients’ experience is what drives clinical practice, often more than measurable changes. Although our population is relatively large, the presence of missing data concerning the use of the *replacement workflow* (AP? category) and the side of worsened symptoms, made an analysis per lead less reliable. For the purpose of data analyses, we focused on worsened outcomes and did not distinguish between unchanged or improved outcomes, because patients are more likely to report worsening than improvement of effect, and because the interest of this study was on the potential issues arising from neurostimulator replacement.

In conclusion, based on our findings, we recommend accounting for possible changes in therapeutic effect following AP replacements by keeping patients under prolonged observation or scheduling (telephone) consultations at short interval, in order to minimize the discomfort and avoid potentially dangerous situations: this is particularly important when the previous stimulator was programmed in CV or with multi-cathode configurations. Patients should be informed of the possibility of an altered clinical response after replacement and advised to contact the center or, when possible, to make adjustments in stimulation amplitude using the patient controller. Finally, we recommend continued use of the currently advised workflow, at least until a revised algorithm will become available.

## Methods

### Study design and patient selection

We conducted a multicenter, retrospective study in six of the seven Dutch DBS centers; Leiden University Medical Center / Haga Teaching Hospital (LUMC / Haga), Amsterdam UMC, University Medical Center Groningen (UMCG), Maastricht University Medical Center (MUMC+), Medisch Spectrum Twente (MST) and Radboud University Medical Center (Radboudumc). Formal approval of the study was waived by the Medical Ethics Committee Leiden The Hague Delft. In four centers, an Opt-Out consent procedure was used, in the other three centers, written informed consent was obtained by the local investigators.

We included all PD patients with STN-DBS, who had an AP RC/PC replacement between February 2020 (the time when Percept™ entered the market in Netherlands) and September 30, 2023, and had stable stimulation settings, which was defined as no change in active contact, pulse width or frequency within the three months preceding replacement.

As a control group, we collected data from an equal number of patients per center who had undergone an AA RC/PC replacement. Selection of patients for the AA group started with the most recent AA replacement and proceeded retrospectively until the number of patients matched that of the AP group at the corresponding center. In addition, data from the most recent previous AA replacement for all patients within the AP group were collected. These “previous AA replacements” were not included in the general AA replacement group.

In order to avoid overrepresentation of a single center, we limited the inclusion to a maximum of 25 patients per group per center, starting by the most recent. For the AA group, the use of the “copy-paste” function for transferring settings was assumed for all IPG replacements. The AP group was subdivided into three groups: 1) replacements in which the *replacement workflow* was appropriately used (AP+); 2) replacements in which the *replacement workflow* was not or inappropriately used (AP−); and 3) replacements in which it was unknown whether the *replacement workflow* was used (AP?).

### General replacement procedure

The IPG replacements were performed by a neurosurgeon, resident or specialized physician assistant in same-day admission as an outpatient surgical procedure. Local or general anesthesia was applied and antibiotic prophylaxis used according to the local protocol. After replacement, the IPG was immediately turned on.

### Data acquisition and outcomes

The therapeutic outcome was defined as the patient’s experience of an improved/unchanged or worsened effect after IPG replacement as reported in the electronic patient file, including whether parameters adjustment up to three months after IPG replacement was needed. All data related to change in therapeutic effect, including change in Parkinson’s symptoms or stimulation-induced side effects, in the first three months after replacement, were collected. Whether a certain symptom was due to reduced stimulation effect (i.e., increased PD symptoms) or increased stimulation effect (i.e., stimulation-induced side effects) was defined by the interpretation of each treating clinician and whether stimulation was increased or decreased to improve symptoms. The centers reported the symptoms as left sided, right sided, axial, or unknown. When symptoms indicated both increased PD symptoms and stimulation side effects in the same patient, axial and/or without specification of side, it was defined as a ‘combination’. In case of uncertainty, two experienced researchers defined the final outcome based on predetermined criteria.

### Data analysis

Data were analyzed using IBM SPSS Statistics, version 29 ^15^ or RStudio 2025.09.2. ^16^. All statistical tests were two-sided with a significance level of 0.05. Baseline patient and DBS characteristics between groups were compared using the Mann-Whitney U or Kruskal-Wallis test for numerical variables, and the chi-square test (using Monte Carlo method (10.000 iterations) in case of small groups) or Fisher’
ss exact test for categorical variables. For baseline DBS characteristics, only data of leads with known therapeutic effect per lead were analyzed.

For the primary analyses, a binary logistic regression was performed using the kind of replacement, with or without use of the *replacement workflow* (AP+, AP– or AA) as the predictor and whether there was a worsened or unchanged/improved therapeutic effect as outcome.

Furthermore, the adjustment of settings after replacement was classified as “Settings adjusted because of worsened effect”, “Settings adjusted because of other reason” and “ No adjustment of settings”. The AP? group was not included in the primary analysis.

For patients with a previous AA replacement, the therapeutic effect of the most recent one was compared to the effect of their AP replacement using a McNemar test. In this analysis, we did include the patients belonging to the AP? group.

For the secondary analyses, we looked at health care utilization, defined as the number of DBS setting adjustments made during contact with a clinician (either by telephone or at the outpatient clinic) after IPG replacement. The adjustments of settings done independently by patients were not included since these did not directly reflect health care utilization. Results regarding the specification of worsened therapeutic effect (increased PD symptoms or stimulation side effects) were compared between groups with a chi-square test (using the Monte Carlo method (10.000 iterations)). Because a substantial proportion of data regarding the side of symptoms was missing, the main analyses were performed per patient, with per-lead analysis conducted only secondarily.

## Data availability

The datasets generated and analyzed during the current study are available from the corresponding author within the informed consent restrictions upon reasonable request.

## Code availability

Not applicable.

## Acknowledgements

We thank all the patients for making their data available for this study. We would like to acknowledge the support of Ms. Angelique Boekee and Mr. Gaetano Leogrande, from Medtronic Inc. for providing technical information on the devices. This study was funded by the HagaOnderzoeksfonds (WF O5/JW/T23-060). The funder played no role in study design, data collection, analysis and interpretation of data, or the writing of this manuscript.

## Author contributions

E.A.M.Y.R.: 1A/B/C, 2A/B, 3A/B

M.F.C.: 1A/B, 2C, 3B

S.v.d.G.: 1A/C, 2C, 3B

B.J.K., M.N.S., M.B., J.M.t.K., S.J.E.V., M.L.K., M.C.T-C, E.v.V., E.M.d.R., R.A.J.E., C.F.E.H., T.A.v.E., N.A.v.d.G., R.Z.: 1C, 2C, 3B

E.W.v.Z.: 2A/C, 3B

1. Research project: A. Conception, B. Organization, C. Execution;
2. Statistical Analysis: A. Design, B. Execution, C. Review and Critique;
3. Manuscript Preparation: A. Writing of the first draft, B. Review and Critique

## Competing interests

T.A.v.E. was supported by the Dutch Research Council ZonMW (VENI grant 09150162410198). M.B. received research funding from the Amsterdam UMC TKI-PPP grant (2021 & 2023 call), the EU Joint Programme – Neurodegenerative Disease Research (JPND) project (2021 call) and stichting ParkinsonFonds (2023 & 2025) and Medtronic (2023 - 2025), all paid to the institution.

M.F.C. is an independent consultant for research and educational issues of Medtronic (all fees to institution), is an independent consultant for research by INBRAIN (all fees to institution), provides research support/contracted research for Boston Scientific (all fees to institution) and received speaking fees for: ECMT (CME activity), and Boston Scientific (all fees to institution).

N.A.v.d.G. provides research support/contracted research and speaking fees for Boston Scientific (all fees to institution).

E.M.d.R. has received travel and accommodation support from Boston Scientific and Abbott for educational events.

All other authors report no competing interests relevant to this study.

## Notes

### Author Declarations

Formal approval of the study was waived by the Medical Ethics Committee Leiden The Hague Delft.

