## Supplementary for "Change in deep brain stimulation effect in Parkinson’s disease after replacement with a new generation neurostimulator"

### Supplementary information

| **Center** | **Screened** | | **Primary inclusions** | | **Included for analysis*** | |
|---|---|---|---|---|---|---|
| | **AP†** | **AA‡** | **AP** | **AA** | **AP** | **AA** |
| Amsterdam UMC | 26§ | 30 | 25 | 25 | 22 | 19 |
| LUMC / Haga | 25 | 25 | 19 | 19 | 18 | 18 |
| UMCG | 16 | 16 | 16 | 16 | 14 | 13 |
| MUMC+ | 11 | 10 | 10 | 10 | 10 | 10 |
| MST | 6 | 7 | 6 | 6 | 6 | 6 |
| Radboudumc | 4 | 4 | 4 | 4 | 4 | 3 |
| ***Total*** | *88* | *92* | *80* | *80* | *74* | *69* |

**Supplementary table 1: number of patients with Parkinson´s disease and STN-DBS considered for inclusion**

Patients were excluded according to predefined exclusion criteria. Groups are divided per participating center and type of IPG replacement. ***** Final inclusions after secondary exclusion. † Total AP in the considered period (the predefined maximum per center was 25), ‡ Amount of screened Activa® to Activa® replacements in order to include the same amount of replacements as in the Activa® to Percept™ group, § Amount of screened Activa® to Percept™ replacements in order to include 25 replacements. AA = Activa® to Activa® replacement, Amsterdam UMC = Amsterdam University Medical Center, AP = Activa® to Percept™ replacement, IPG= implantable pulse generator, LUMC / Haga = Leiden University Medical Center / Haga Teaching Hospital, MST = Medisch Spectrum Twente, MUMC = Maastricht University Medical Center, Radboudumc = Radboud University Medical Center, UMCG = University Medical Center Groningen.

| **Parameter (n=patients)** | **AA (n=69)** | **AA with unknown effect (n=11)** | **P-value** | **AP (n=74)** | **AP with unknown effect (n=3)** | **P-value** | **P-value AA vs AP** |
|---|---|---|---|---|---|---|---|
| Male, n (%) | 47 (68.1) | 7 (63.6) | 0.742 | 54 (73.0) | 2 (66.7) | 1.000 | 0.584 |
| Age years, median (IQR) | 65.0 (61.5-72.0) | 69.0 (64.0-73.0) | 0.142 | 68.0 (62.8-73.0) | 73.0* | 0.123 | 0.149 |
| Disease duration years, median (IQR) | 17.0 (13.3-21.8)† | 19.0 (13.0-22.0) | 0.739 | 18.5 (15.0-22.3) | 25.0* | 0.277 | 0.131 |
| Duration DBS-treatment years, median (IQR) | 7.0 (5.0-10.0) | 6.0 (5.0-15.0) | 0.632 | 8.0 (6.0-11.0) | 7.0* | 0.644 | **0.048** |
| **Parameter (n=leads)** | **AA (n=108)** | **AA with unknown effect (n=22)** | **P-value** | **AP (n=124)** | **AP with unknown effect (n=6)** | **P-value** | **P-value AA vs AP** |
| Type of lead | | | | | | | |
| 3389, n (%) | 104 (96.3) € | 22 (100.0.) | 1.000 | 118 (95.2) ‡ | 6 (100.0) | N/A | 0.223 |
| 3387, n (%) | 2 (1.9) € | 0 (0.0) | | 0 (00) ‡ | 0 (0.0) | | |
| Stimulation configuration | | | | | | | |
| CC, n (%) | 14 (13.0) | 2 (9.1) | 1.000 ¶ | 16 (12.9) | 0 (0.0) | 1.000 ¶ | 1.000 ¶ |
| Monopolar (CC), n (%) | 12 (11.1) | 2 (9.1) | | 12 (9.7) | 0 (0.0) | | |
| Bipolar (CC), n (%) | 2 (1.9) | 0 (0.0) | | 4 (3.2) | 0 (0.0) | | |
| CV, n (%) | 94 (87.0) | 20 (90.9) | | 108 (87.1) | 6 (100.0) | | |
| Monopolar (CV), n (%) | 53 (49.1) | 16 (72.7) | | 76 (61.3) | 6 (100) | | |
| Bipolar (CV), n (%) | 20 (18.5) | 4 (18.2) | | 17 (13.7) | 0 (0.0) | | |
| Double monopolar (CV), n (%) | 9 (8.3) | 0 (0.0) | | 11 (8.9) | 0 (0.0) | | |
| Asym. double bip. (CV), n (%) | 5 (4.6) | 0 (0.0) | | 2 (1.6) | 0 (0.0) | | |
| Interleaved (CV), n (%) | 7 (6.6) | 0 (0.0) | | 2 (1.6) | 0 (0.0) | | |
| Therapeutic current (mA), median (IQR) | 3.5 (2.6-4.2) § | 2.6 (2.0-3.7) # | **0.049** | 3.7 (2.7-4.2) ** | 3.3 (2.9-3.9) | 0.755 | 0.830 |
| Frequency (Hz), median (IQR) | 130 (125-180) | 130 (130-135) | 0.799 | 130 (130-180) | 130 (130-160) | 0.963 | 0.579 |
| Pulse width (μs), median (IQR), range | 60 (60-60), 60-120 | 60 (60-60), 60-110 | 0.514 | 60 (60-60), 40-120 | 60 (60-60), 60-60 | 0.536 | 0.139 |

**Supplementary table 2. Patient characteristics per group (in- as well as secondarily excluded) and DBS characteristics per lead on Activa® IPG before replacement**

For the analysis per lead between the total AA and AP group, only data of patients with known therapeutic effect per lead were included: 108/138 AA, 124/148 AP**.** * = IQR not reported due to small sample size. † Available for 68/69 patients, € Available for 106 leads, ‡ Available for 118 leads, ¶ Result for distribution of total CC and CV between groups, § Available for 79 leads, # Available for 12 leads, ** Available for 94 leads. AA= Activa® to Activa® replacements, AP = Activa® to Percept™ replacements, IPG= implantable pulse generator.

| Cases in which the *replacement workflow* was not or inappropriately used (AP- replacements) | Amount of patients n=16 (%) |
|---|---|
| Workflow not used | 11 (68.8) |
| Voltage to current 1 on 1 | 4 (25.0) |
| Started with lower amplitude; | |
| and adjusted according to symptoms | 3 (18.8) |
| because of dyskinesia in the face prior to replacement | 1 (6.3) |
| Total Electrical Energy Delivered (TEED) | 1 (6.3) |
| Started with different (one higher, one lower) amplitude, for unknown reason | 2 (12.5) |
| Workflow inappropriately used (misunderstanding for double monopolar) | 5 (31.3) |

**Supplementary table 3: Cases in which the *replacement workflow* was not/inappropriately used after Activa® to Percept™ replacements**

AP- replacements = Activa® to Percept™ replacement not/inappropriately using workflow.

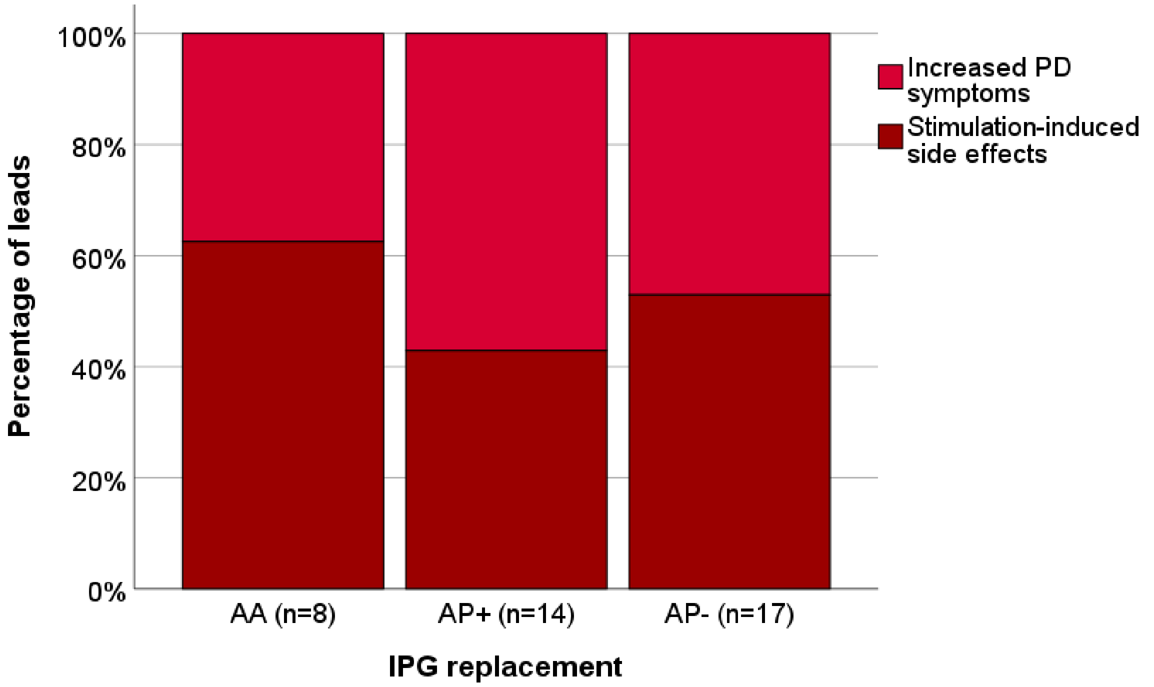


**Supplementary figure 1: Specification of worsened therapeutic effect per lead after IPG replacement**

Replacements divided in AA (Activa® to Activa® replacement), AP+ (Activa® to Percept™ replacement using workflow) and AP– (Activa® to Percept™ replacement not/inappropriately using workflow) with specification of worsened therapeutic effect: increased PD symptoms (understimulation, light red); stimulation-induced side effects (overstimulation, dark red). Leads with known specified worsened therapeutic effect: AA 8/28, AP+ 14/28, AP– 17/21. This accounts for less than the total number of leads for all worsened patients because some patients had one sided worsening of effect (2 AA, 4 AP+, 3 AP– ). PD= Parkinson´s disease, IPG= implantable pulse generator.

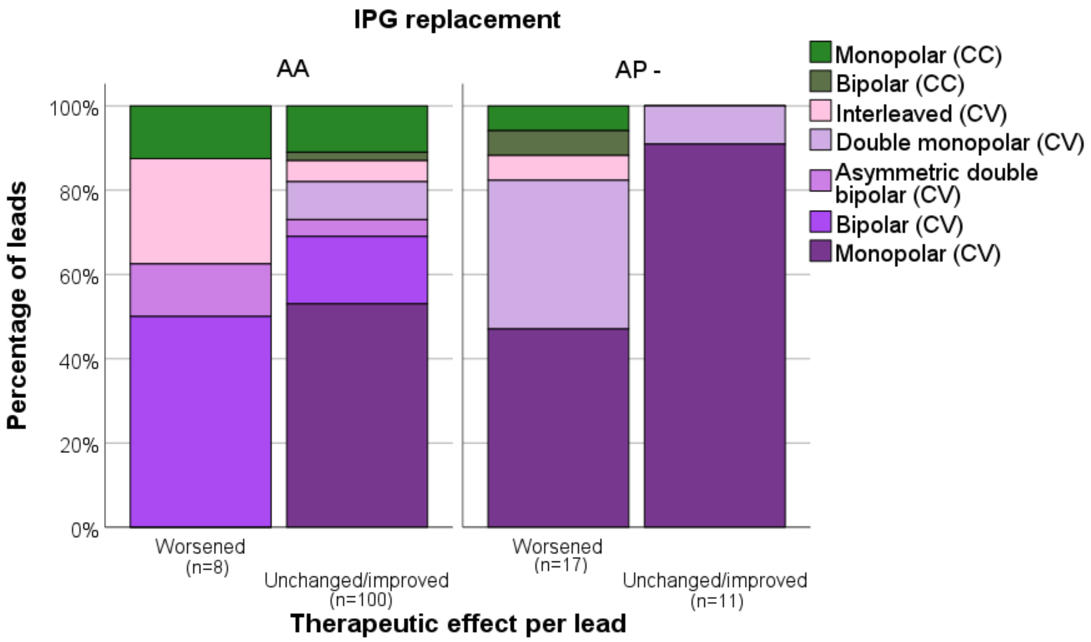


**Supplementary figure 2: Specification of pre-replacement stimulation configuration per lead for Activa® Activa® and Activa® Percept™ – replacements**

Replacements divided AA (Activa® to Activa® replacement) and AP– (Activa® to Percept™ replacement not/inappropriately using workflow) in worsened and unchanged/improved therapeutic effect. Leads with known therapeutic effect: AA: 108/138 (For the 5 patients with improved therapeutic effect, the effect per lead (n=10) was not known, thus not represented in the graph); AP–: 28/32(one patient with improved therapeutic effect). CC = constant current, CV = constant voltage, IPG= implantable pulse generator.
